# Analytical sensitivity and effectiveness of different SARS-CoV-2 testing options

**DOI:** 10.1101/2021.11.26.21265946

**Authors:** Nico Lelie, Marco Koppelman, Harry van Drimmelen, Sylvia Bruisten

## Abstract

We prepared severe acute respiratory syndrome-coronavirus-2 (SARS-CoV-2) working standards and reference panels from a pool of swab fluid samples before and after inactivation by beta-propiolactone and quantified viral load in nucleic acid amplification technology (NAT) detectable RNA copies/mL using limiting dilution analysis. The following 50% lower limits of detection (LOD) were estimated by probit analysis as compared to detection limits of rapid antigen tests on 1.5 fold dilutions of the native material: Roche cobas PCR 1.8 (1.0-3.3), Hologic Aptima TMA 6.6 (4.4-9.9), DRW SAMBA 15 (7-30), Molgen LAMP 23 (13-42), Fluorecare antigen 50,000, Abbott Panbio antigen 75,000 and Roche antigen 100,000 copies/mL. One 50% Tissue Culture Infectious Dose (TCID_50_)/mL of culture fluid was estimated to be equivalent to approximately 1000 RNA copies/mL (2700-4300 International Units) in our working standard. When assuming this level as start of contagiousness in a log-linear ramp up viremia model with 10-fold rise of viral load per day for the B.1 (Wuhan) type we estimated relative time points of first detectability of early infection by the different SARS-CoV-2 assays from the LODs mentioned above. The four NAT assays would be able to detect early viremia 40-66 hours earlier than the 1000 copies/mL infectivity threshold, whereas the three antigen tests would become positive 41-48 hours later. Our modeling of analytical sensitivity data was found to be compatible with clinical sensitivity data of rapid antigen tests and confirms that NAT assays are more reliable than antigen assays for identifying early infected asymptomatic individuals who are potentially infectious.

## 1 INTRODUCTION

Several studies compared the clinical sensitivity of molecular assays and rapid antigen tests on nasopharyngeal or nasal swab samples for detection of severe acute respiratory syndrome-coronavirus 2 (SARS-CoV-2) infection^1-6^. The limitation of these comparison studies is that separate swabs must be taken for detection of SARS-CoV-2 RNA and antigen respectively, whereas these individuals were usually tested because of (mild) symptoms and already had higher viral loads. Only limited data is available from studies that directly compared the analytical sensitivity of different nucleic acid amplification technology (NAT) systems and lateral flow devices for rapid detection of SARS-CoV-2 antigen using serial dilutions of swab fluid with a known viral load^6-8^. In this report we used reference preparations from pooled swab samples in Viral Transport Medium (VTM) before and after inactivation by beta-propiolactone^9,10^ for comparison of the analytical sensitivity of different SARS-CoV-2 assays. These working standards were first quantified in NAT detectable SARS-CoV-2 RNA copies/mL using limiting dilution analysis and later - at the time the World Health Organization (WHO) International Standard became available - also in International Units (IU/mL). Additionally, by comparison of NAT detection limits on USA-WA1-2020 culture fluid^11^ the amount of NAT detectable RNA copies per 50% Tissue Culture Infectious Dose (TCID_50_) was estimated.

Since the rapid antigen tests are known to miss a considerable proportion of infected individuals^1,2^ we also evaluated two NAT methods that were developed for fast detection of SARS-CoV-2 infection i.e. a loop-mediated isothermal amplification (LAMP) assay and a so called Simple amplification based assay (SAMBA)^12^ and compared their sensitivity with two widely used NAT systems as reference methods i.e. a real time polymerase chain reaction (PCR) assay^11^ and a transcription mediated amplification (TMA) assay^13^. By testing dilution series of our native and inactivated working standards the lower limits of detection (LOD) of these NAT systems were compared with those of three rapid antigen assays. We then used the analytical sensitivity data, including the impact of testing minipools of six samples (MP6) using the PCR assay^11^, to evaluate the effectiveness of these different SARS-CoV-2 detection options in identifying infected individuals during the early asymptomatic phase of infection. For this evaluation a log-linear ramp up viremia model was assumed using best estimates for the viral doubling time and start of potential contagiousness^14,15^.

## 2 METHODS

### 2.1 Preparation of native and inactivated SARS-CoV-2 standards and reference panels

Working standards were prepared from a pool of remnant fluid of swab samples, kindly provided by the Public Health Laboratory (GGD) of Amsterdam. Remnant samples were fully anonymized before use. The study procedure was evaluated by the Medical Ethics Committee of the Academic Medical Center in Amsterdam (W21_507 # 21.559) and deemed not to require a full review of the board. Swab samples in Viral Transport (GLY) Medium were collected in August to November 2020 at the time only the B.1 (Wuhan) type of SARS-CoV-2 virus circulated in the Netherlands. Samples were known to be tested positive using the Hologic TMA test with RLU values >1200. Approximately 90 mL of this pooled medium constituted the native SARS-CoV-2 standard. Seventy-five mL of this native pool was inactivated with 0.14% beta-propiolactone for 5 hours at 23°C, followed by 18 hours incubation at 2-8°C^10,11^, to produce the inactivated standard. This inactivated standard was used for preparing a 10 member (3 and 10-fold) dilution panel in a 2% plasma solution in phosphate buffered saline (PBS), which was used for replicate testing by different NAT methods to determine the 95% and 50% LODs by probit analysis. In addition a 20 member panel composed of 1.5-fold dilutions of both the native and inactivated standard was prepared for comparing the analytical sensitivity of different rapid antigen assays. The standards and reference panels were prepared by BioQControl (Heiloo, The Netherlands) via gravimetrically recorded dilution steps and were snap frozen in liquid nitrogen before storage at −80°C.

### 2.2 Quantification of SARS-CoV-2 RNA and antigen in working standards before and after inactivation

Ten-fold standard dilutions of both the native and inactivated SARS-CoV-2 standards were made in 2% plasma-PBS and tested in duplicate in the cobas PCR assay (Roche Molecular Systems) to compare the SARS CoV-2 RNA Ct values for both the ORFa/b and E gene targets. The yield after inactivation was determined by parallel line analysis on Ct values for the two PCR targets separately. A panel composed of 3 and 10-fold dilutions of the inactivated standard was tested in multiple replicate tests in different NAT methods to allow for estimation of the limit of detection (LOD) by probit analysis. The concentration in NAT detectable RNA copies/mL that was assigned to the inactivated standard was based on the 63% LOD in the most sensitive NAT method. We assumed that this assay reached 100% NAT efficiency or a 63% LOD of 1 detectable RNA copy per amplification reaction as follows from Poisson distribution in limiting dilution analysis. The concentration in RNA copies/mL in the inactivated working standard was then calculated by correcting for the input volume in the NAT method. In addition, 3 and 10-fold dilutions of the inactivated standard were tested in duplicate against similar concentrations of the WHO International Standard 20/146 in the cobas PCR assay. The conversion factor between NAT detectable RNA copies and International Units (IUs) was calculated by parallel line analysis on Ct values for both ORFa/b and E gene targets in the cobas assay. The concentration in the native standard was derived from the value assigned to the inactivated standard and the measured yield after inactivation, whereby the average was taken for the two PCR targets in the cobas assay. The yield of SARS-CoV-2 antigen after inactivation was determined by comparison of geometric mean detection endpoint titers on 1.5-fold dilutions of both the native and inactivated standard in three rapid antigen tests.

### 2.3 Estimation of SARS-CoV-2 viral load in culture fluid of known infectivity

We compared the 95% LOD expressed in TCID_50_/mL on USA-WA1-2020 culture fluid reported in the package insert of the Roche cobas assay^11^ with the 95% LOD expressed in NAT detectable RNA copies/mL assigned to our inactivated working standard to estimate the dose of SARS-CoV-2 RNA copies per TCID_50_.

### 2.4 SARS-CoV-2 assays evaluated for analytical sensitivity

To determine the 95% and 50% LOD by probit analysis the inactivated SARS-CoV-2 standard dilution panel with estimated viral RNA concentrations varying between 33,784 and 1.1 copies/mL was tested in multiple replicates by the following NAT assays: Roche cobas PCR^11^, Hologic Aptima TMA^13^, LAMP [prototype assay, Molgen, Veenendaal, the Netherlands] and Diagnostics of the Real World (DRW) SAMBA II assay^12^. The 1.5 fold SARS-CoV-2 standard dilution panel with concentrations varying between 800,000 and 60,000 copies/mL of both the native and inactivated standards was tested in duplicate by three rapid antigen lateral flow devices, i.e. Panbio COVID-19 Ag Rapid Test (Abbott Rapid Diagnostics Jena GmbH), SARS-CoV-2 Rapid Antigen Test (Roche Diagnostics GmbH) and Fluorecare COVID-19 Spike Protein Test Kit (Shenzhen Microprofit Biotech Co, Ltd, kindly provided by Sander Brus, PreVViral, Amsterdam, The Netherlands). The latter Chinese assay exclusively targeted the spike protein, while the other kits targeted the nucleocapsid protein. For each of the antigen tests the panel members were mixed 1:1 with the sample buffer from the kits and the required volume was added to the reaction hole of the lateral flow devices. The panel members that gave a clearly visible line were scored as + whereas the next panel member(s) showing a faint line was recorded as ± or indeterminate. The viral concentration in the 1:1 mixed sample buffer with the last + and ± reactivity were recorded as endpoint titers, whereas the input copy numbers in the volumes added to the reaction holes were recorded as well. If the duplicate reactions were discrepant (+ and ± or ± and -) the series with the lowest endpoint titers were taken as final positive (+) and indeterminate (±) result.

### 2.5 Statistics

The 95%, 63% and 50% LODs on the BioQ SARS-CoV-2 analytical sensitivity panel were calculated by probit analysis and relative sensitivities of assays by parallel line probit analysis using SPSS software. The potency of the inactivated SARS-CoV-2 standards against the native standard and the WHO International Standard 20/146 was calculated from the Ct values in the cobas assay using parallel line analysis.

### 2.6 Assumptions for modeling assay conversion times during early infection

We assumed a log-linear ramp-up viremia model in which the viral load of the B.1 (Wuhan) type increased on average approximately 10-fold per day (mean viral doubling time 7.2 hours) during the initial phase of infection and on average 10,000-fold per day (mean viral doubling time 1.8 hours) with the currently circulating B.1.617.2 (delta) variant according to observations of Li et al during two outbreaks in Guangdong province in China^14^. Although these Chinese investigators observed a considerable variation in viral growth curves between individual cases during daily quantitative PCR testing they estimated a 1000-fold higher mean viral load at the day of PCR conversion with the delta variant than with 19A/19B strains during an earlier outbreak in the beginning of 2020.

The 50% LOD NAT conversion point of the most sensitive assay was arbitrarily set at time point zero and the 50% LOD NAT conversion points found with (the most sensitive targets of) the other assays, including a MP6 PCR option^11^, were calculated using the assumptions for the viral growth curves mentioned above. The rapid antigen detection limits with a faint reaction line (±) on the 1.5 fold native standard dilutions were used to calculate the time intervals between the 50% NAT conversion points and the rapid antigen test conversion points.

## 3 RESULTS

### 3.1 Quantification of SARS-CoV-2 RNA and antigen in working standard before and after inactivation

Three 10-fold dilutions of SARS-CoV-2 standards before and after inactivation were tested in duplicate in the cobas SARS-CoV-2 assay and Ct values for the two PCR gene targets are shown in Supplemental Table 1. With parallel line assay the distance between Ct values (95% confidence interval (CI)) before and after inactivation was 1.96 (1.79-2.12) for ORFa/b and 2.67 (2.57-2.75) for E gene targets respectively. The potency (95% CI) of the native standard was thus estimated to be 2^1.96^ = 3.88 (3.46-4.35) fold higher than the inactivated standard based on ORFa/b gene PCR and 2^2.67^ =6.34 (5.97-6.79) fold higher based on E gene PCR (on average a 4.96-fold difference). Hence, the recovery after treatment of the pool of swab fluid with beta-propiolactone was 25.8 (23.0-28.9)% for the ORFa/b gene and 15.7 (14.8-16.8)% for the E gene (yield on average 20.2%).

From the proportions of reactive tests in three different NAT assays on inactivated standard dilutions the 63% LOD (and 95% confidence interval (CI)) was calculated by parallel line probit analysis and the results are shown in Table 1. We assumed that the NAT method with the highest sensitivity (the cobas assay for ORFa/b target) reached 100% NAT efficiency or a 63% LOD of 1 NAT detectable RNA copy per amplification reaction as follows from Poisson distribution statistics. Since for each replicate cobas PCR test a volume of 400 uL was used as input in the amplification reaction the concentration at the 63% LOD was thus set at 2.5 copies/mL for the cobas PCR ORFa/b assay. Therefore initially assigned concentrations to the standard dilution panel (data not shown) needed to be adjusted with a recalibration factor and from the dilution factors we calculated that the viral load in the undiluted inactivated standard was 3.38 × 10^6^ NAT detectable RNA copies/mL and 4.96-fold higher (see above) in the native standard (1.68 × 10^7^ copies/mL). With these assigned RNA copies/mL to the SARS-CoV-2 standards the calculated NAT efficiency was 100 (53-187)% and 54 (24-94)% in the cobas ORFa/b and E PCR tests respectively, whereas it would be 67 (42-102)% in the Aptima TMA assay and 40 (41-73)% in the LAMP assay (Table 1).

**Table 1.** Estimation of SARS-CoV-2 RNA concentration in inactivated standard based on Poisson distribution assuming 100% NAT efficiency of most sensitive assay

| Assay | volume amplified/<br>input volume in assay<br>μL | n<sup>^</sup> | Recalibrated<br>63% LOD<br>copies/mL<br>(95% CI) | relative<br>sensitivity<br>factor<br>(95% CI) | NAT efficiency<br>(95% CI)# |
| --- | --- | --- | --- | --- | --- |
| cobas PCR<br>target 1 [ORFa/b] | 400/400<br>μL | 8 | 2.50<br>(1.33-4.73) | 1.00<br>(reference) | 100%<br>(53-187)% |
| cobas PCR<br>target 2 [E] | 400/400<br>μL | 8 | 4.67<br>(2.66-8.52) | 0.53<br>(0.31-0.93) | 54%<br>(29-94)% |
| Aptima TMA | 167/500<br>μL | 14 | 8.95<br>(5.88-14.2) | 0.28<br>(0.19-0.42) | 67%<br>(42-102)% |
| Prototype LAMP | 80/400<br>μL | 6 | 30.9<br>(17.0-60.4) | 0.08<br>(0.04-0.15) | 40%<br>(21-73)% |

<sup>^</sup> number of replicates tested per member of P0356 SARS CoV-2 standard dilution panel

When comparing the dilutional titers in three rapid antigen tests on the 1.5-fold dilution panel we estimated that the SARS-CoV-2 antigen concentration was 1.8 (range 1.3-2.5) fold lower in the beta-propiolactone-inactivated standard than the untreated standard, whereas the RNA concentration was 5.0 (3.9-6.3)-fold lower after inactivation. Hence, the antigenicity relative to the RNA concentration was found to be 2.7 (range 2.5-3.0) fold higher in the inactivated standard than in the native standard.

### 3.2 Calibration of inactivated SARS-CoV-2 working standard against the WHO International Standard

For calibration of the inactivated SARS-CoV-2 standard against the WHO 20/146 standard we tested dilution series in the cobas SARS-CoV-2 assay in duplicate (Supplemental Table 2a) and analyzed the Ct values using parallel line analysis with exception of the lowest concentration to improve parallelism. The potency difference or the amount of IUs per NAT detectable RNA copy was 4.29 (4.44-5.36) for the ORFa/b target and 2.68 (2.19-3.29) for the E gene target (Supplemental Table 2b).

### 3.3 Relation between viral load and infectivity in culture fluid

The 95% LOD (with 95% confidence interval (CI)) in the package insert of the cobas assay was estimated in TCID_50_/mL using culture fluid of the SARS-CoV-2 Strain USA-WA1-2020. For the ORFa/b gene the Roche package insert reported a 95% (CI) of 0.007 (0.005-0.036) TCID_50_/mL, whereas on our working standard we estimated the 95% LOD at 8.3 (4.5-18.6) copies/mL for this PCR gene target. According to this comparison 1 TCID_50_ would be equivalent to 1186 (847-6098) PCR detectable RNA copies in our working standard.

### 3.4 Analytical sensitivity of different NAT and rapid antigen assays

Table 2 gives the proportion of reactive results and mean measurement values on the inactivated standard dilutions for the two most sensitive NAT assays, i.e. the cobas real time PCR and Aptima TMA assay. Similar data are also available for the LAMP and SAMBA assays (data not shown). Table 3 shows the reactivity of three rapid antigen tests on 1.5 fold serial dilutions of the native and inactivated working standards. In Table 4 we combined the analytical sensitivity data of the four NAT systems and three rapid antigen assays. The cobas assay for ORFa/b was the most sensitive with 50% and 95% LOD of 1.8 (1.0-3.3) and 8.3 (4.5-18.6) copies/mL if used on single samples and 11.1 (5.9-20.1) and 49.8 (27-112) if used on minipools of 6 samples. In parallel line probit analysis the Aptima TMA assay was 3.6 (2.4-5.3) fold less sensitive than cobas PCR with 50% and 95% LODs of 6.6 (4.4-9.9) and 29.7 (18.4-60.1) copies/mL. The LAMP assay was 12.4 (6.7-25) fold less sensitive than the Roche assay with estimated 50% and 95% LODs of 23 (13-42) and 102 (54-251) copies/mL. The SAMBA II assay was evaluated later using the same reference panel and in a separate probit analysis the 50% LOD and 95% LOD were 15 (7-30) and 133 (44-447) copies/mL respectively.

**Table 2.**
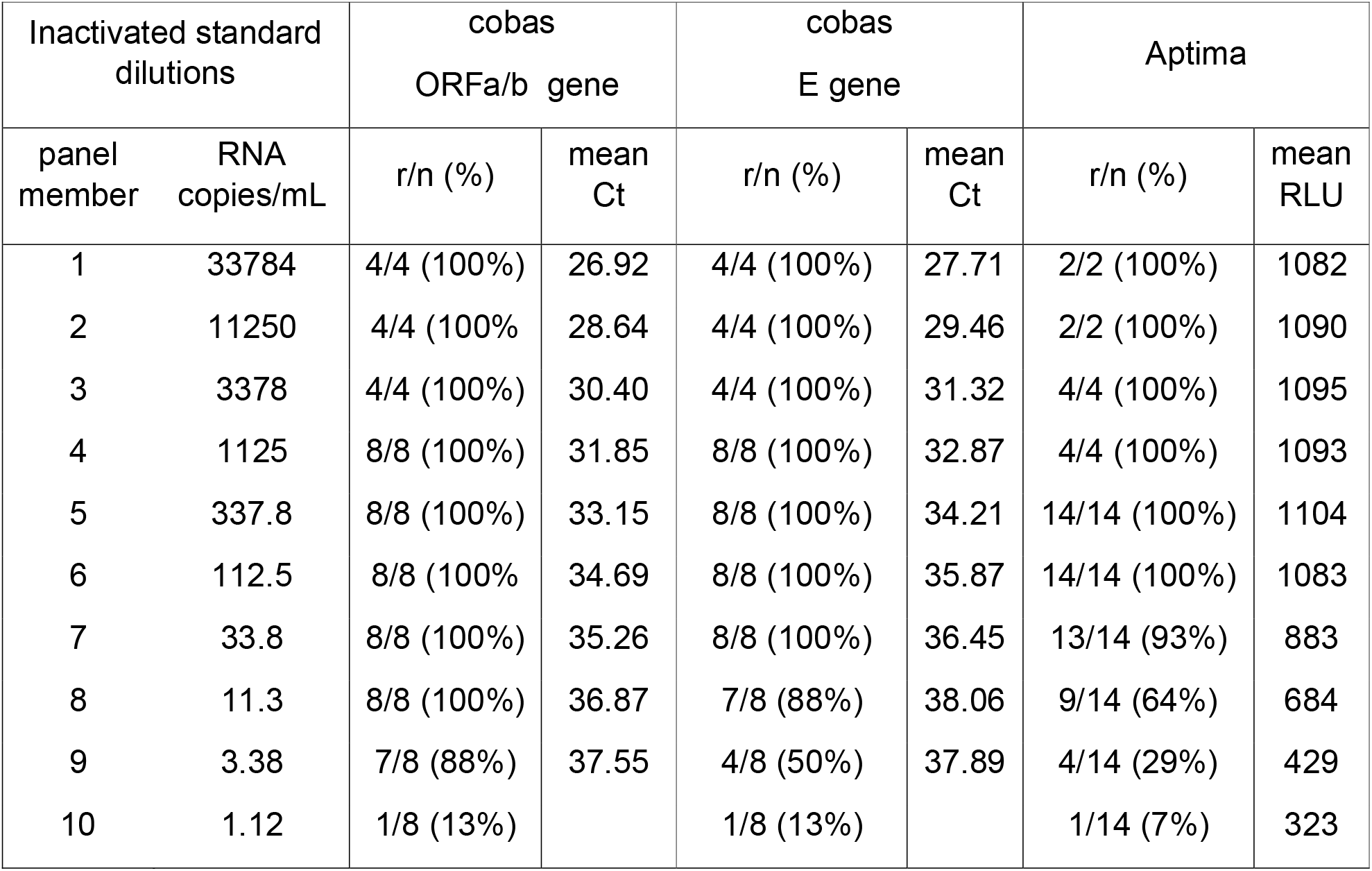
Proportion reactive and average assay response values on an inactivated SARS-CoV-2 standard dilution panel in two NAT systems

| Inactivated standard dilutions |  | cobas<br>ORFa/b gene |  | cobas<br>E gene |  | Aptima |  |
| --- | --- | --- | --- | --- | --- | --- | --- |
| panel member | RNA copies/mL | r/n (%) | mean Ct | r/n (%) | mean Ct | r/n (%) | mean RLU |
| 1 | 33784 | 4/4 (100%) | 26.92 | 4/4 (100%) | 27.71 | 2/2 (100%) | 1082 |
| 2 | 11250 | 4/4 (100%) | 28.64 | 4/4 (100%) | 29.46 | 2/2 (100%) | 1090 |
| 3 | 3378 | 4/4 (100%) | 30.40 | 4/4 (100%) | 31.32 | 4/4 (100%) | 1095 |
| 4 | 1125 | 8/8 (100%) | 31.85 | 8/8 (100%) | 32.87 | 4/4 (100%) | 1093 |
| 5 | 337.8 | 8/8 (100%) | 33.15 | 8/8 (100%) | 34.21 | 14/14 (100%) | 1104 |
| 6 | 112.5 | 8/8 (100%) | 34.69 | 8/8 (100%) | 35.87 | 14/14 (100%) | 1083 |
| 7 | 33.8 | 8/8 (100%) | 35.26 | 8/8 (100%) | 36.45 | 13/14 (93%) | 883 |
| 8 | 11.3 | 8/8 (100%) | 36.87 | 7/8 (88%) | 38.06 | 9/14 (64%) | 684 |
| 9 | 3.38 | 7/8 (88%) | 37.55 | 4/8 (50%) | 37.89 | 4/14 (29%) | 429 |
| 10 | 1.12 | 1/8 (13%) |  | 1/8 (13%) |  | 1/14 (7%) | 323 |

**Table 3.**
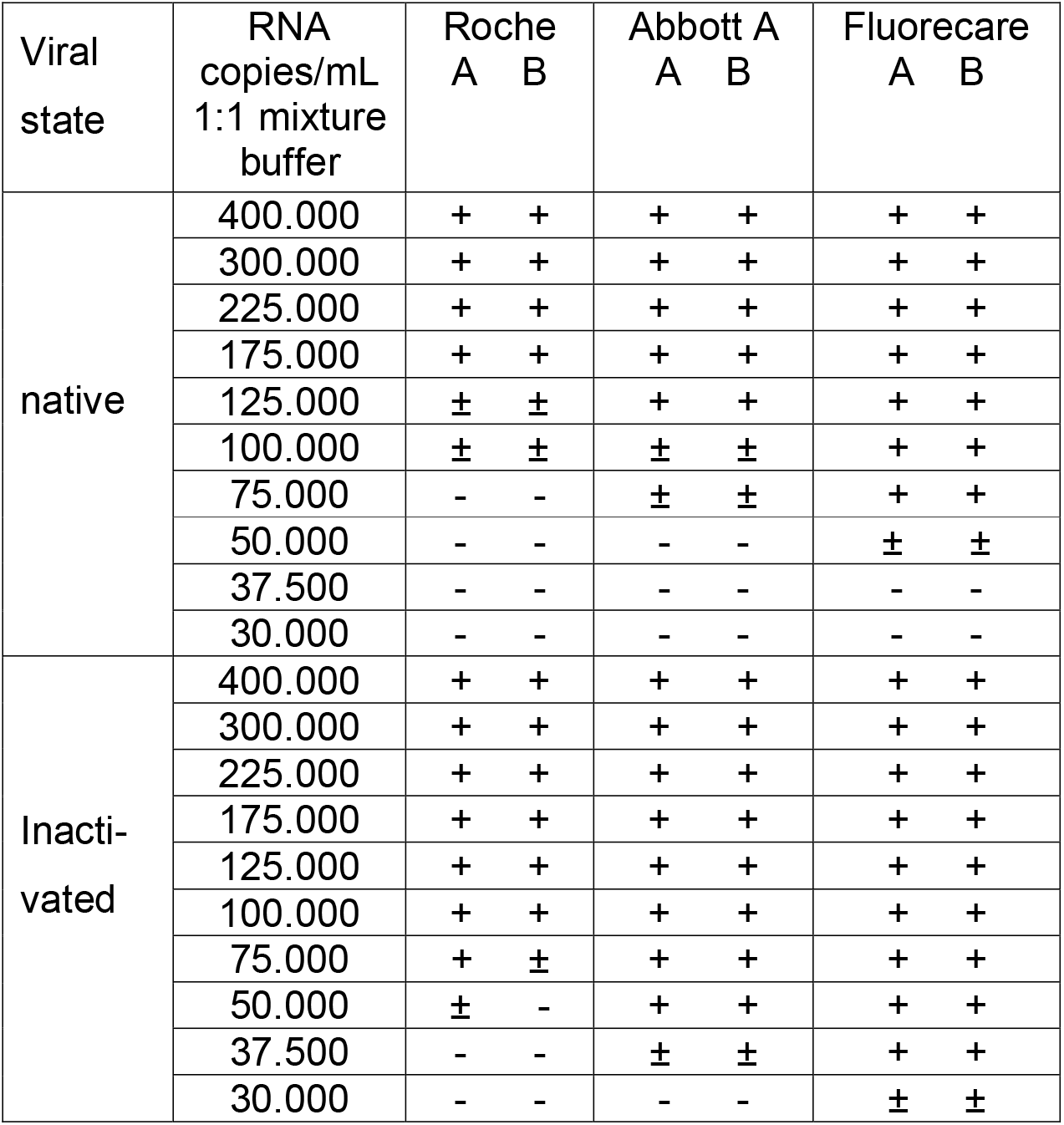
Comparison of analytical sensitivity of three rapid antigen tests on 1.5 fold dilutions of SARS-CoV-2 working standards before and after inactivation

**Table 4.** Comparison of LODs of different SARS-CoV-2 NAT and antigen assays and relative sensitivity factors on working standards before and after inactivation

| Test method | Viral state of standard | Replicate tests per dilution | LOD reactivity | RNA copies/mL (95%CI) | Relative insensitivity factor □ |
| --- | --- | --- | --- | --- | --- |
| Roche cobas PCR ORFa/b | inactivated | 8 | 50%<br>95% | 1.8 (1.0-3.3)<br>8.3 (4.5-18.6) | 1<br>1 |
| Roche cobas PCR E gene | inactivated | 8 | 50%<br>95% | 3.5 (2.0-6.0)<br>15.5 (8.7-34.6) | 1.9<br>1.9 |
| Roche MP6# PCR ORFa/b | inactivated | 8 | 50%<br>95% | 11.1 (5.9-20.1)<br>49.8 (27-112) | 6.0<br>6.0 |
| Hologic Aptima TMA | inactivated | 14 | 50%<br>95% | 6.6 (4.4-9.9)<br>29.7 (18.4-60.1) | 3.6<br>3.6 |
| Molgen LAMP | inactivated | 6 | 50%<br>95% | 22.8 (12.7-42.1)<br>102.5 (54.1-251) | 12.4<br>12.4 |
| DRW SAMBA ORF | inactivated | 8 | 50%<br>95% | 15.0 (7.1-30.1)<br>133 (44.3-447) | 8.1<br>16.0 |
| DRW SAMBA N | inactivated | 8 | 50%<br>95% | 150 (79.2-271)<br>597 (315-4168) | 81.1<br>71.9 |
| Roche Antigen | inactivated | 2 | ±<br>+ | 75000<br>100000 | 41667<br>12048 |
| Abbott Antigen | inactivated | 2 | ±<br>+ | 37500<br>50000 | 20833<br>6024 |
| Fluorecare Antigen | inactivated | 2 | ±<br>+ | 30000<br>37500 | 16667<br>4518 |
| Roche Antigen | native | 2 | ±<br>+ | 100000<br>175000 | 55556<br>21084 |
| Abbott Antigen | native | 2 | ±<br>+ | 75000<br>125000 | 41667<br>15060 |
| Fluorecare Antigen | native | 2 | ±<br>+ | 50000<br>75000 | 27778<br>9036 |

□ according to comparison of 50% and 95% LODs respectively for NAT methods as calculated by probit analysis. For antigen assays detection endpoint titers with indeterminate (±) reactivity were compared with the 50% LOD in cobas PCR, whereas endpoint titers with positive (+) reactivity were compared with the 95% LOD in cobas PCR

The analytical sensitivity of the rapid antigen assays on 1.5-fold dilutions of the native standard dilutions were 28,000 to 56,000 fold lower than the cobas PCR assay when comparing the lowest concentration giving indeterminate antigen reactivity with the 50% LOD in PCR, whereas the LODS with rapid antigen test positive reactivity were 9,000 to 21,000-fold higher than the 95% LOD in the cobas assay (Table 4). Interestingly the Chinese Fluorecare assay targeting the spike protein was slightly more sensitive than the Abbott and Roche antigen assays targeting the nucleocapsid protein.

### 3.5 Modeling of SARS-CoV-2 assay conversion time points during early infection

When assuming a log-linear ramp up viremia model with a daily 10-fold increase of viral load for the B.1 (Wuhan) type virus and 10,000 fold per day for the B.1.617.2 (delta) variant (see methods) the time intervals between assay conversion during early infection were estimated from the 50% LODs for the NAT options and the LODs with indeterminate (±) reactivity for the antigen assays (Figure and Supplementary Table 3). Assuming an infectivity threshold at 1 TCID_50_/mL or around 1000 copies/mL in swab fluid (see above) the 50% NAT conversion points were estimated to be 40-66 hours earlier for the B.1 (Wuhan) strain. By contrast, according to our model the rapid antigen assays would become positive 41-48 hours later than this infectivity threshold and therefore have a higher probability to miss contagious individuals in an early phase of infection than NAT assays. If the assumption of on average 1000-fold faster replication of the delta variant is correct^14,15^ the time intervals between assay conversion points become four-fold shorter (Figure, Supplementary Table 3).

## 4 DISCUSSION

In the present study we directly compared the analytical sensitivity of four different NAT assays and three lateral flow devices for rapid SARS CoV-2 antigen detection on dilution series of a pool of swab fluid samples before and after inactivation by beta-propiolactone. The viral load in the native and inactivated material was quantified in NAT detectable RNA copies/mL by limiting dilution analysis and in IU/mL against the WHO International Standard. Moreover our working standards were used to calculate viral load in culture fluid of known infectivity and we estimated that 1 TCID_50_ was equivalent to approximately 1000 NAT detectable RNA copies in our working standard. We were surprised that the antigen concentration relative to the RNA concentration was found to be 2.7 (range 2.5-3.0) fold higher in the inactivated standard than in the native standard. We speculate that the beta-propiolactone treatment has destroyed or modified subgenomic RNA fragments from human cells that were present in the pool of swab GLY samples, but had less impact on full length RNA genomes packaged in virions and on the antigenicity of the nucleocapsid or spike protein. For discussion on further details of the standardization in RNA copies or IUs in swab fluid before and after inactivation and the relation to TCID_50_ we refer to the Appendix.

On 1.5 fold dilutions of the native working standard the rapid antigen tests were found to be 28,000 to 56,000 fold less sensitive than the 50% LOD of 1.8 (1.0-3.3) copies/mL achieved by the most sensitive NAT assay in our comparison study, which was the cobas PCR assay. The Aptima TMA assay was 3.6 fold less sensitive than this PCR assay, whereas the SAMBA and LAMP assays were 8-12 fold less sensitive.

Assuming a ramp up viremia model of 1 Log10 per day^14,15^ we estimated that the Fluorecare, Abbott and Roche antigen assays were able to detect early viremia 4-5 days later than the Roche cobas and Hologic Aptima NAT assays, whereas the cobas PCR in MP6 format, SAMBA and LAMP would be approximately 0.5-1 day slower in detecting early viremia than the most sensitive NAT options. More importantly, we estimated that the NAT assays were able to detect early viremia 40-66 hours before the estimated tissue culture infectivity conversion point at approximately 1000 copies/mL in the ramp up phase of viremia, whereas the antigen assays became positive 41-48 hours later.

Rapid antigen tests have long been promoted as an effective tool in preventing SARS-CoV-2 transmission since they detected the majority of PCR positive symptomatic individuals and it was assumed that the probability of contagiousness was highest in antigen positive individuals^1,2^. It is possible that the airborne transmissibility of SARS-CoV-2 between humans in close contact starts at a viral load that is lower than the estimated tissue culture infectivity limit of 1000 copies/mL in swab fluid. Taking considerable uncertainty ranges in the exact conversion time points of assays and the start of contagiousness into account we believe that our modeling of analytical sensitivity data helps to understand what the effectiveness is of using different SARS-CoV-2 test options in timely identifying potentially contagious subjects. Our modeling of the time points of assay conversion during early viremia matches with evaluation data of laminar flow antigen detection methods in large clinical studies in the Netherlands^1,2^. One of these studies^2^ found a clinical sensitivity of 59% with two widely used rapid antigen tests when asymptomatic individuals were tested 5 days after potential exposure, whereas a higher proportion of 73-84% of the exposed subjects with symptoms were antigen reactive. These proportions are compatible with our model of early dynamics of viremia and estimated antigen conversion points as shown in the Figure.

The calculated time points of SARS-CoV-2 RNA and antigen conversion in the different assays were based on best estimates of the viral growth curve before the B.1.17 (alpha) and B.1.617.2 (delta) variants circulated in the Netherlands. Currently the delta variant is the dominant strain and therefore the modeled NAT and antigen conversion time points in the Figure were adjusted for the more rapid replication of this variant causing a 1000-fold higher rise in viral load per day^14,15^. If this assumption is correct the assay conversion points and infectivity levels in ramp up phase of viremia would be reached 4 times earlier by the delta variant than by the original wildtype virus. Since viral loads of the delta variant in vaccine breakthrough infections were found to be comparable to those in unvaccinated individuals^16,17^ a model of 10,000-fold increase of viral load per day may be generalizable to all early infected individuals by the delta variant regardless of the vaccination status. One can imagine that the effectiveness of testing and isolation of infected individuals during current outbreaks of the delta variant is even more dependent on the sensitivity of the assay and the rapid turnaround time of the test results than in the past with the original virus.

We therefore recommend the use of NAT methods rather than antigen assays for more reliable identification of contagious individuals. In a fast NAT screening setting where one receives the test results not immediately at the testing site but a few hours later by e-mail, one could just as well opt for a laboratory that uses a highly sensitive NAT method and receive the test result within the same day. Finally - if the infection rate in the target population is <4% one could also make use of more cost-effective NAT testing of pooled swab samples as recently demonstrated for MP4 using the SAMBA assay^18^ and shown in the present report for MP6 using the cobas PCR assay.

In conclusion, we estimated up to 56,000 fold differences in detection limits between several SARS-CoV-2 detection methods on a pool of swab fluid samples before inactivation. Translating these analytical sensitivity data to assay conversion time points using a ramp up viremia model helped to better understand what the effectiveness is of these testing options in detecting early (asymptomatic) infection in potentially contagious individuals.

## Data Availability

All data produced in the present work are contained in the manuscript

---

**Figure.**
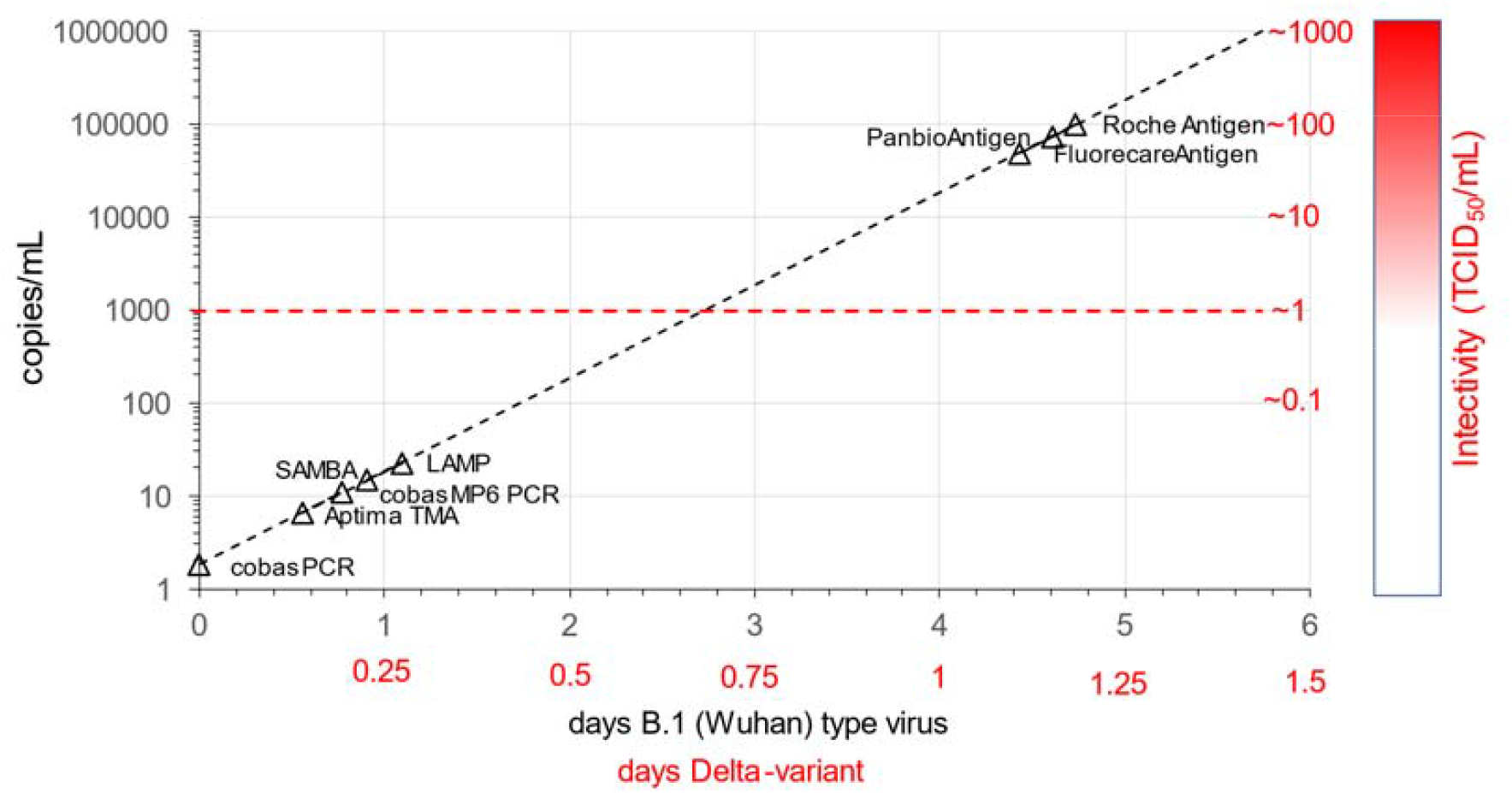
Estimated time point of conversion of different NAT and rapid antigen detection options during ramp up phase of SARS-CoV-2 viremia in relation to potential infectivity level of swab fluid in tissue culture. The mean viral doubling time (with wide confidence limits) was estimated at 7.2 hour based on viral growth curves of the original B.1 (Wuhan) type virus and estimated to be 1.8 hour with the currently circulating B.1.617.2 (delta) variant^14,15^.

