## Supplemental Tables for "Analytical sensitivity and effectiveness of different SARS-CoV-2 testing options"

**Supplemental Table 1.** Ct values in Roche cobas SARS-CoV-2 RNA test on standard dilutions before and after inactivation by beta-propiolactone.

| Standard | | A-series | | B-series | |
|---|---|---|---|---|---|
| | | ORFa/b | E gene | ORFa/b | E gene |
| Viral state | Dilution | Ct | Ct | Ct | Ct |
| native | 10 | 21.95 | 22.10 | 21.78 | 21.96 |
| native | 100 | 25.05 | 25.13 | 24.91 | 25.18 |
| native | 1000 | 27.87 | 28.20 | 28.09 | 28.32 |
| inactivated | 10 | 23.54 | 24.55 | 23.77 | 24.67 |
| inactivated | 100 | 26.93 | 27.87 | 26.95 | 27.86 |
| inactivated | 1000 | 30.15 | 30.99 | 30.04 | 30.95 |

**Supplemental Table 2a.** Ct values on WHO and BioQ SARS-CoV-2 standard dilution series in cobas PCR assay

| | series A | | series B | |
|---|---|---|---|---|
| WHO 20/146 IU/mL | ORF Ct1 | E Ct2 | ORF Ct1 | E Ct2 |
| 100000 | 27.63 | 27.78 | 27.31 | 27.38 |
| 30000 | 29.16 | 29.26 | 29.43 | 29.61 |
| 10000 | 30.71 | 30.82 | 30.39 | 30.61 |
| 3000 | 32.20 | 32.61 | 33.07 | 33.05 |
| 1000 | 32.94 | 33.37 | 33.27 | 33.77 |
| BioQ inactivated RNA copies/mL | ORF Ct1 | E Ct2 | ORF Ct1 | E Ct2 |
| 33784 | 26.94 | 27.75 | 26.54 | 27.30 |
| 11249 | 28.74 | 29.60 | 28.78 | 29.68 |
| 3378 | 30.34 | 31.25 | 30.09 | 31.02 |
| 1125 | 31.75 | 32.75 | 31.92 | 32.56 |
| 338 | 33.17 | 34.03 | 33.26 | 34.21 |

**Supplemental Table 2b**. Potency of BioQ inactivated standard against WHO 20/146 standard in cobas SARS-CoV-2 PCR assay

| Target | parameter | Value (95% CI) |
|---|---|---|
| ORF | delta Ct | 2.10 (1.78-2.42) |
| | IU/copy | 4.29 (3.44-5.36) |
| E | delta Ct | 1.42 (1.13-1.72) |
| | IU/copy | 2.68 (2.19-3.29) |

**Supplemental Table 3.** Estimated assay conversion times relative to the most sensitive assay, i.e. cobas PCR (Supplemental table 3a) and relative to an assumed infectivity threshold of 1000 NAT detectable RNA copies/mL equivalent to 1 $TCID_{50}$ (Supplemental table 3b) during ramp up phase of viremia of B.1 (Wuhan) type SARS-CoV-2 virus and the B.1.617.2 (delta) variant assuming a 10-fold and 10,000 fold daily rise of viral load respectively.

Supplemental table 3a

| assay | 50% LOD copies/mL^ | B.1 (Wuhan) type | | B1.617.2 (delta) variant |
|---|---|---|---|---|
| | | days | hours | hours |
| cobas PCR | 1.8 | 0 | 0 | 0 |
| Aptima TMA | 6.6 | 0.56 | 13.4 | 3.3 |
| cobas MP6 PCR | 11.1 | 0.78 | 18.7 | 4.7 |
| SAMBA | 15.0 | 0.91 | 21.8 | 5.4 |
| LAMP | 22.8 | 1.09 | 26.2 | 6.6 |
| 1 $TCID_{50}$/mL | 1000 | 2.74 | 65.9 | 16.5 |
| Fluorecare antigen | 50000 | 4.44 | 106.5 | 26.6 |
| Panbio antigen | 75000 | 4.61 | 110.7 | 27.7 |
| Roche antigen | 100000 | 4.74 | 113.7 | 28.4 |

^50% NAT LODs or ± antigen LODs on native standard before inactivation

Supplemental table 3b

| assay | 50% LOD copies/mL^ | B.1 (Wuhan) type | | B.1.617.2 (delta) variant |
|---|---|---|---|---|
| | | days | hours | hours |
| cobas PCR | 1.8 | -2.74 | -65.9 | -16.5 |
| Aptima TMA | 6.6 | -2.19 | -52.5 | -13.1 |
| cobas MP6 PCR | 11.1 | -1.97 | -47.2 | -11.8 |
| SAMBA | 15.0 | -1.84 | -44.1 | -11.0 |
| LAMP | 22.8 | -1.65 | -39.6 | -9.9 |
| 1 $TCID_{50}$/mL | 1000 | 0.00 | 0.0 | 0.0 |
| Fluorecare antigen | 50000 | 1.69 | 40.6 | 10.2 |
| Panbio antigen | 75000 | 1.87 | 44.8 | 11.2 |
| Roche antigen | 100000 | 1.99 | 47.8 | 12.0 |

^50% NAT LODs or ± antigen LODs on native standard before inactivation
