## Appendix for "Analytical sensitivity and effectiveness of different SARS-CoV-2 testing options"

### Standardization details

In this standardization study we compared LODs of different NAT methods with the detection endpoint titers of rapid antigen tests on 1.5 fold dilutions of a pool of swab fluid samples before and after inactivation by beta-propiolactone. These working standards were quantified in NAT detectable RNA copies/mL by limiting dilution analysis and in IU/mL by comparison against the WHO 20/146 standard, whereby one NAT detectable RNA copy by the cobas PCR assay in the inactivated working standard was found to be equivalent to 4.29 (4.44-5.36) IUs for the ORFa/b target and 2.68 (2.19-3.29) IUs for the E gene.  Hence the IU/copy conversion factors in the cobas PCR assay were found to be 1.60 (1.57-1.63) higher for the ORFa/b gene than for the E gene. Similarly we found that  the amount of E gene targets was reduced 1.64 (1.55-1.72) fold more than the amount of ORFa/b gene targets by treatment of the working standard with beta-propiolactone. We speculate that this slight but significant difference in recovery is caused by presence of unequal amounts of subgenomic RNA of the ORFa/b and E genes derived from human cells that were present in the swab fluid pool before inactivation. Another explanation – although less likely – is that beta-propiolactone renders more ORFa/b than E gene targets undetectable in the cobas PCR assay.  It may be that the inactivation of the cell culture-derived WHO International Standard by acid and heat treatment acts differently than the chemical inactivation of our working standard. However the relative amount of detectable genomic and subgenomic RNA of the ORFa/b and E genes in the WHO standard preparation before inactivation is unknown.  It must be noted that the difference in Ct value between target 1 [ORF] and 2 [E] on the USA-WA1-2020 culture fluid used for determining analytical sensitivity in the Roche cobas package insert was on average 2.42 as compared to a difference of 0.92 on our inactivated standard. In terms of a potency (or relative detectability) this is a factor 5.37 versus 1.88 for the two standards. This suggests that there is still 2.8 fold more of E gene RNA in our inactivated working standard than in the cultured USA-WA1-2020 culture fluid. Hence there are significant differences in the potency of ORFa/b and E genes in different standards according to quantification in the cobas SARS-CoV-2 assay. Our calibration in RNA copies/mL was based on assuming 100 (53-187)% NAT efficiency of the ORFa/b gene in the Roche cobas assay and 67 (42-102)% in the Hologic Aptima test. If the amount of IUs would be equal to the true amount of RNA molecules the NAT efficiency of the two assays would be 23 (12-44)% and 16 (10-24)% respectively.

When comparing the relationship between viral load and infectivity in WA1-2020 culture fluid we estimated that 1 $TCID_{50}$ would be equivalent to 1186 (847-6098) RNA copies of the ORFa/b gene in our working standard. According to the certificate of analysis NR-52281 (BEI resources) 1 $TCID_{50}$ of the USA WA1-2020 strain would be equivalent to 7393 RNA copies according to quantification in Droplet Digital PCR. Our estimate was also not compatible with the 95% positive tissue culture infectivity limit of $2.17 \times 10^5$ copies/mL in clinical studies [1,2] according to an in house calibration curve of the Erasmus University Medical Center (EUMC) [1]. This limit corresponded with a Roche cobas E gene Ct value ≤30 [1]. A Ct value of 25 corresponded with $4.87 \times 10^6$ E gene copies/mL according to EUMC [1], which in our calibration in PCR detectable RNA copies/mL was 26-fold lower and corresponded with a concentration of 186,000 RNA copies/mL. Hence, the EUMC 95% positive culture limit of $2.17 \times 10^5$ EUMC copies/mL may be equivalent to approximately 8300 copies/mL according to our calibration, which level seemed compatible with our estimate of a 50% tissue culture infectivity limit of approximately 1000 PCR detectable RNA copies/mL.

We were surprised that the antigen concentration relative to the RNA concentration was found to be 2.7 (2.5-3.0) fold higher in the inactivated standard than in the native standard. We speculate that the beta-propiolactone treatment has destroyed or modified subgenomic RNA fragments from human cells that were present in the pool of swab GLY samples, but had less impact on full length RNA genomes packaged in virions and on the antigenicity of the nucleocapsid or spike protein. Additionally, antigen epitopes hidden in immune complexes may have been released by beta-propiolactone, although this seems less likely since the virus in the GLY-pool must have been dominated by swab samples with very high viral load from antibody negative (or low reactive) individuals.

In conclusion, there seem to be significant differences between standards in the amount of SARS-CoV-2 RNA copies and IUs for different NAT gene targets, which balance is also affected by inactivation. Therefore assay detection limits as well as the minimum infectious dose in tissue culture are dependent on the reference preparation used for calibration in RNA copies.
